# The Impact of Buoy on Hydration Status of Active Men and Women

**DOI:** 10.1101/2024.09.28.24314547

**Authors:** Cary Boyd-Shiwarski, Evan Ray, Harikesh Subramanian, Nicole Zharichenko, Amy Monroe, Aman Mahajan

**Affiliations:** University of Pittsburgh, Department of Medicine, Renal and Electrolyte Division; Department of Anesthesiology and Perioperative Medicine, University of Pittsburgh, University of Pittsburgh Medical Center (UPMC), Pittsburgh, PA

**Keywords:** Electrolytes, Renal, Supplement, Dehydration, Body Hydration Index

## Abstract

**Background:** Hypo-hydration is a major health concern that affects performance and is associated with increasing morbidity and growing health care costs. There is an emerging interest in optimum hydration and identifying how factors such as ingestion rate and beverage composition affect hydration. This study examined three beverages with varying ingestion rates and measured markers of hydration.

**Methods:** Thirty healthy, active participants between ages 18-45 years were given three different beverages on three separate days. The beverages were of identical volumes (1 Liter), but differed in the rate of ingestion, carbohydrate content and electrolyte content. Beverage 1 (Buoy, San Diego, CA) and water-alone were both consumed at a metered rate of one liter over four hours, whereas Beverage 2 was used as a positive control and consumed at a bolus rate of one liter in 30 minutes.

**Findings:** After six hours Beverage 1 significantly improved markers of hydration compared to water-alone or Beverage 2. Beverage 1 decreased cumulative urine output vs water-alone by 32% (absolute difference -0.33L; CI ± -0.16 to -0.51) and vs Beverage 2 by 26% (absolute difference - 0.26L; CI ± -0.13 to -0.38). Beverage 1 increased the beverage hydration index vs water-alone by 64% (absolute difference +0.64L; CI ± 0.36 to 0.92) and vs Beverage 2 by 48% (absolute difference +0.53L; CI ± 0.30 to 0.76)

**Interpretation:** Beverage 1 is superior to water-alone at improving hydration when ingested at similar rates. Moreover, metered ingestion of Beverage 1 improved hydration compared to a bolus ingestion of Beverage 2, this could be due to the dissimilar ingestion rates and/or beverage composition.

**RESEARCH IN CONTEXT:** Despite the overwhelming number of commercial hydration beverages on the market, there are only a very limited number of studies that address whether these beverages are actually effective at improving hydration. Using PubMed and Google Scholar using the search term “Beverage Hydration Index” with the search date from 2016-2024 (2016 was when the Beverage Hydration Index was established) we found less than 10 articles on this topic that used the beverage hydration index to assess the efficacy of popular beverages and supplements, and none of them have previously evaluated the efficacy of Beverage 1 (Buoy). Additionally, only one other study assessed how that rate of beverage ingestion can influence the beverage hydration index. This current study has found Beverage 1 increased the beverage hydration index vs water-alone by 64% (absolute difference +0.64L; CI ± 0.36 to 0.92). We propose that Beverage 1 increases the beverage hydration index due to its abundance of electrolytes including sodium and chloride, as it does not contain carbohydrates, protein, or artificial sweeteners that are common in other commercial hydration beverages. Identifying beverages that improve hydration compared to water-alone can play an important role in preventing severe hypohydration and dehydration, including renal failure, seizures, arrythmia, and altered mental status. It has been estimated that over half a million hospitalizations per year are due to dehydration with a cost of over 5.5 billion United States dollars(1). Thus, there are both clinical and economic reasons to identify simple, cost-effective methods to promote euhydration.

## INTRODUCTION

Preserving hydration through renal handling of water and electrolytes is pivotal to survival(2–4). Euhydration refers to a state of “normal” body water content (with changes less than ±0.2-0.5% of total body water), with “hypo- and hyperhydration” being either a water deficit or excess beyond these limits—and the term “dehydration” refers to a 3% or greater loss of total body water (3, 5, 6). Severe hypohydration, defined as a body water deficit greater than normal daily fluctuation, can result in renal failure, arrhythmia, fainting, headache, fatigue, seizures and mental status changes (7). Humans can only survive for approximately three days without water due to ongoing losses through insensible loss, sweating, and excretion. A major challenge for maintaining euhydration is compensating for ongoing losses with appropriate intermittent water and electrolyte intake(8).

Studies have shown that during exercise humans rarely ingest enough fluid to match their sweat loss, with some studies suggesting fluid intake usually only compensates for 50% of sweat loss during physical activity(9–12). Hypohydration can have detrimental effects on cognition, technical skill, and physical performance(4, 13); and conversely increasing fluid ingestion during exercise improves core body temperature, heart rate and stroke volume(14). However, athletes are not the only population vulnerable to hydration deficits. Those that work in hot environments, the elderly, those with increased GI output (i.e. vomiting, diarrhea, ostomy output, inflammatory bowel disease), poor oral intake, pregnancies complicated with hyperemesis gravidarum, peri-operative patients, and patients with long term health conditions including cancer, diabetes and alcohol use can all be affected by hypo-hydration. It has been estimated that over half a million hospitalizations per year are due to dehydration with a cost of over 5.5 billion United States dollars(1). Thus, there are both practical and clinical reasons to identify simple, cost-effective methods to promote euhydration over longer periods of time.

The primary method for maintaining euhydration is by consuming beverages which can account for 80% of daily water intake(4, 15). The beverage hydration index (BHI) is a method developed by Maughan *et al*., to compare the short-term hydration efficacy of different beverages(8). The BHI compares the hydration potential of a given beverage to that of plain water, with water having a BHI of 1.0. Beverages that promote greater diuresis than water will have a BHI <1.0 and are assumed to be ineffective at promoting oral rehydration, whereas beverages that limit diuresis compared to water will have a BHI > 1.0 and are assumed to be effective at promoting oral rehydration(15).

Buoy® (San Diego, CA) is a natural sourced, organic compliant, Food and Drug Administration (FDA) compliant, unflavored, sugar and sweetener free, electrolyte supplement, primarily made up of sodium, chloride and potassium with 87 other trace minerals, that can be added dropwise to any beverage to increase the beverages’ electrolyte content. Buoy is designed to be consumed in small aliquots in beverages throughout the day and has been purported to prevent hypohydration and electrolyte loss. This study is a comparison between Buoy Hydration drops (from here on referred to as Beverage 1), water, and another commercially available hydration product (Beverage 2, Nuun Sport, Seattle, WA) to determine if the claims of improved hydration status were accurate. Beverage 2 was used as a positive control as it was previously shown to improve markers for hydration(16). The primary aim of this study was to determine whether Beverage 1 improved hydration compared to water alone, and the secondary aim was to measure whether there were any hydration differences between Beverage 1 and Beverage 2.

## METHODS

### Ethics Approval

This is a single-center prospective, cross-over, placebo-controlled clinical trial at the University of Pittsburgh Medical Center (UPMC). All experimental procedures were approved by the University of Pittsburgh Medical Center Institutional Review Board approval number STUDY22090018 and consent was approved for eligible patients. The trial was registered at www.clinicaltrial.gov under NCT05768789. Participants completed a health history and informed consent prior to the start of the study.

### Study Population

This study enrolled adult participants who were healthy volunteers who could walk up at least one flight of stairs without difficultly. The inclusion criteria included any male or female patient ≥18 to 45 years of age, non-tobacco users, negative pregnancy test in women of childbearing potential, creatinine ≤ 1.2 milligram/decilitre (mg/dL), no known underlying medical conditions, willingness to refrain from alcohol for 24 hours prior to testing day, willingness to refrain from strenuous exercise for 24 hours prior to each test day, without any obvious signs or symptoms of infection. Patients who were excluded, had a creatinine lab value > 1.2 mg/dL, proteinuria / hematuria / glucosuria based on urine dipstick, diagnosed medical condition that would impede results (congestive heart failure, hypertension, coronary artery disease, chronic kidney disease, history of electrolyte abnormality), pregnancy, use of diuretics within two weeks prior to first day of trial, active infection based on symptoms (bacterial or viral), hemodynamic abnormality at screening visit with blood pressure less than 100/60 millimetres of mercury (mmHg) or greater than 140/90 mmHg. Prior to initiation of the study, participants completed informed consent and a health history. During the initial visit female participants were administered a pregnancy test.

### Experimental Protocol

The protocol was devised based on methods from prior publications(8, 15–17). The subjects completed three study visits on separate days. Testing visit 1 was for Beverage 1 (Buoy Hydration Drops). Testing visit 2 was for water-alone. Testing visit 3 was for Beverage 2 (Nuun Sport, Seattle, WA). The type of bottled water used (for reconstitution as well as control arm) in all three visits was Kirkland ® (Brentwood, TN). Participants refrained from vigorous exercise within 24 hours of the study visit. Participants were asked to fast for 10 hours through the night prior to each testing day. Upon waking they were asked to empty their bowel and bladder. If desired, they were allowed to consume one 8-ounce cup of coffee or other clear liquid, with the expectation that the participant would be consistent with its consumption with each testing visit.

During each testing visit, subjects presented between 0700-0800 hours at which time they were asked to empty their bladder again. After resting for five minutes, baseline vitals were taken, including blood pressure, heart rate, weight, bioimpedance (Omron, Kyoto, Japan). Urinalysis dipstick for protein/blood/glucose (Siemens, Berlin, Germany) and i-STAT (Abbott, Lake County, Illinois) measurement for creatinine and electrolytes were done on Visit 1 to confirm eligibility. For Visit 2 and Visit 3, the participants were re-interviewed to review any changes in their medical history that would warrant an additional baseline creatinine and blood/protein test to confirm eligibility.

All arms of the study were repeated in every subject. The first arm during the first study visit was Beverage 1, followed by the second study visit for water-alone (control), and finally the third study visit for Beverage 2.

The total volume of fluid consumed during each of the three study visits was capped at one litre (1L) to minimize the risk of water intoxication(4). The Beverage 1 arm and water-only arm had an identical rate of consumption. For these arms, the entire 1L fluid was administered in a metered fashion over 4 hours. This rate of consumption was chosen to reflect the label recommendation of Beverage 1, to drink the solution in small aliquots throughout the day. The water-only arm followed an identical administration protocol to serve as a true control for Beverage 1. For Beverage 2, the third arm, administration was in a bolus fashion of 1L over 30 minutes, based on prior studies showing that bolus consumption of Beverage 2 significantly improved the hydration status at two and four hours compared to water-alone(16). Thus, the Beverage 2 arm served as the comparison of the current stranded.

Urine was collected at four specific timepoints during the intervention (60, 120, 240, 360 minutes) and the mass of the urine was recorded. If participants needed to micturate between scheduled collection times, the urine that was collected was recorded and combined with the urine collection of the following timepoint. These urine samples were measured and sent to the University of Pittsburgh Medical Center hospital laboratory to be tested for the following electrolytes: sodium (Na+), potassium (K+), chloride (Cl-) and urine osmolarity. Urine creatinine was also tested at these timepoints. Bio-impedance was measured after each urine collection. No other food or beverage was consumed by the participants during the 6 hours of each study visits. After completion of all three portions of the study, participants were compensated $300.00 after completion of three visits. See Figure 1 for protocol timeline.

**Figure 1:**
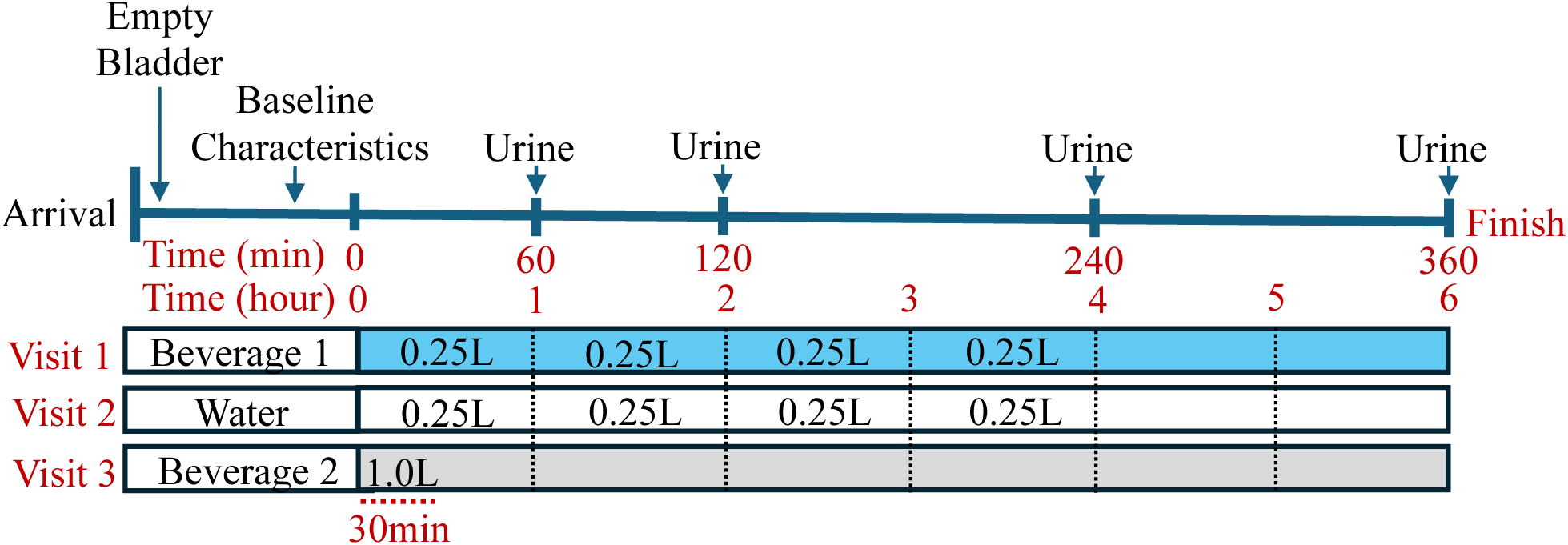
Graphic representing the protocol timeline for each experiment. For Visit 1 (Beverage 1) and Visit 2 (Water-alone) each subject consumed 6.25% of the total volume of one litre (1L) every 15 minutes for four hours. For Visit 3 (Beverage 2) participants consumed 1L within the first 30 minutes. At time 0, the participant began drinking per the above protocol. Urine was collected at 60, 120, 240, and 360 minutes. Abbreviations: min = minute. L = litre.

#### VISIT 1---BEVERAGE 1 INTERVENTION

Each subject consumed Beverage 1 (Buoy Hydration Drops) at a dose containing 600mg/Litre (L) of Na+ over 4 hours while measuring urine output over 6 hours. For Beverage 1, the label recommended dosage is 1.5 millilitres (ml) (50 mg sodium per serving) in multiple servings throughout the day. Therefore, to safely achieve a total dose 600 mg Na+/L (12-fold increase from single dose) we used 18 ml (1.5 ml x 12) of Beverage 1 per 1L of water consumed. This is equivalent to two servings of the extra-strength Buoy Rescue Drops. Each subject consumed 6.25% of the total volume every 15 minutes for four hours. At time 0, the participant began drinking per the above protocol. Urine was collected at 60, 120, 240, and 360 minutes. Bioimpedance was measured after each urine collection.

#### VISIT 2—WATER-ALONE CONTROL

Each subjected ingested 1L of water at a rate of 6.25% of the calculated amount of water every 15 minutes for four hours. At time 0, the participant began drinking per the above protocol. Urine was collected at 60, 120, 240, and 360 minutes. Bioimpedence was measured after each urine collection.

#### VISIT 3—BEVERAGE 2 INTERVENTION

For Beverage 2, each subject consumed 1L of water with two dissolved Nuun Sport Hydration tabs (Nuun, Seattle, WA), containing 600 mg of sodium, over 30 minutes (2 equal volumes every 15 minutes) while measuring urine output over 6 hours (rate of consumption was based on previously published data by Pence *et al.*(16)). At time 0, the participants began drinking per the above protocol and were done drinking at 30 minutes. Urine was collected at 60, 120, 240, and 360 minutes. Bioimpedence was measured after each urine collection. A water-only control with a similar rate of 1L in 30 minutes was not performed as it was previously published(16).

### Calculations

Net fluid balance was calculated as the amount of fluid ingested minus the cumulative urine output at each time point. Participants were considered to be in positive fluid balance if the net fluid balance was > 0. The beverage hydration index (BHI) was developed in 2016 to define the hydration response to any given beverage(8). It assumes that the cumulative urine volume over a fixed period of time denotes the AUC for renal water excretion. Thus, the cumulative urine volume while ingesting water-alone divided by the cumulative urine volume after ingesting the intervention beverage equals the BHI of a beverage. BHI was calculated at each timepoint for Beverage 1 compared to water since equal rates of Beverage 1 and water-alone were consumed at each timepoint. For the BHI comparison of Beverage 1 to Beverage 2, since the beverages were consumed at different rates the BHI was only calculated at a final timepoint of 360 minutes.

### Statistical Analysis

Data were analyzed using Prism software version 10 (GraphPad Software LLC) and presented as either mean ± standard deviation (SD) or mean ± 95% confidence interval (CI) of the mean. Comparisons between two groups were determined by a Student’s T-test. Comparisons between multiple groups and variables were determine using one- or two-way analysis of variance (ANOVA), followed by the appropriate post hoc test, as indicated. P values :: 0.05 were considered statistically significant. The final sample size was determined a priori using G*Power 3.1 based on previous findings(8, 15, 17). Assuming an effect size f^2^(V) = 0.25, α = 0.05; β = 0.2; power = 0.8, requires n = 27 participants per beverage.

## RESULTS

### Overview

Thirty participants successfully completed the study (Table 1), 14 males and 16 females. None of these participants reported a problem with beverage consumption, and all beverages were well-tolerated. No patients were excluded from the analysis. The composition of each beverage is in Table 2.

**Table 1:** Baseline Physical Characteristics of Participants at 1^st^ visit.

| | Male ± SD<br>(n = 14) | Female ± SD<br>(n = 16) | Combined ± SD<br>(n = 30) |
| --- | --- | --- | --- |
| **Age (years)** | 31.6 (± 6.6) | 28.3 (± 4.3) | 29.9 (± 5.6) |
| **Body Weight (kg)** | 88.96 (± 12.8) | 76.9 (± 18.5)* | 82.6 (± 16.9) |
| **BMI** | 27.1 (± 3.8) | 28.1 (± 7.6) | 27.7 (± 6.1) |
| **Body Fat %** | 23.4 (± 8.6) | 33.7 (± 9.3)** | 28.7 (± 10.2) |
| **Skeletal Muscle %** | 36.6 (± 6.0) | 28.6 (± 4.8)*** | 32.5 (± 6.7) |
| **Resting Metabolism (kcal)** | 1868 (± 166) | 1535 (± 199)*** | 1695 (± 248) |
| **Visceral Fat** | 7.4 (± 3.5) | 6.5 (± 2.8) | 7.0 (± 3.2) |
| **Systolic Blood Pressure** | 120.4 (± 3.4) | 119.7 (± 2.9) | 120.0 (± 3.1) |
| **Diastolic Blood Pressure** | 77.1 (± 2.0) | 77.3 (± 1.8) | 77.2 (± 1.9) |
| **Heart Rate** | 77.9 (± 7.5) | 80.0 (± 11.3) | 79.0 (± 9.6) |
| **Serum Creatinine** | 0.94 (± 0.09) | 0.97 (± 0.12) | 0.95 (± 0.11) |
| Statistical significance between male and female was determined using two-tailed T test. Sex was self-reported. *P ≤ 0.05, **P ≤ 0.01, ***P ≤ 0.01. | | | |

**Table 2:** Beverage Composition.

|  | Water | Beverage 1 | Beverage 2 |
| --- | --- | --- | --- |
| **Total Volume** | 1L | 1L | 1L |
| **Sodium (mg)** | Negligible | 600 | 600 |
| **Potassium (mg)** | Negligible | 120 | 300 |
| **Chloride (mg)** | Negligible | 960 | 80 |
| **Magnesium (mg)** | Negligible | 6 | 50 |
| **Calcium (mg)** | Negligible | 6 | 26 |
| **Calories (kcal/L)** | 0 | 0 | 30 |
| **Sugars (g)** | 0 | 0 | 2 |
| **Carbohydrates** | 0 | 0 | 8 |
| Based on manufacture's nutrition labels<br>Beverage 1: 18ml was added to 1 L of water<br>Beverage 2: 2 tabs were added to 1L of water |  |  |  |

### Baseline Physical Characteristics

Baseline characteristics are shown in Table 1, including age, weight, BMI, body composition, blood pressure and heart rate. Several factors were influenced by sex including body weight, body composition (fat versus muscle), and resting metabolism. Factors there were not significantly different between males and females include age, BMI, blood pressure, heart rate, and baseline kidney function.

### Beverage 1 (Buoy) significantly decreased urine volume

Two-way ANOVA revealed both a time-effect (P <0.0001) and beverage-effect (P <0.0001), and a time x beverage effect (P < 0.0001). The rate of urine output (L/minute) reflected the differences in the rate of ingestion (Figure 2a). Both Beverage 1 and water-alone had a slow “metered” rate of ingestion (1L/240 minutes) and urine volume peaked at 240 minutes and then began to decline. Whereas Beverage 2 had a rapid “bolus” rate of ingestion (1L/30 minutes) that was reflected by larger urine volume during the first 120 minutes that rapidly declined by 240 minutes (Figure 2a). Prior studies have shown that when Beverage 2 and water-alone were ingested at a similarly rapid rate (1L/30 minutes), the rate of urine output was similar at 60 minutes, 180 minutes and 240 minutes, and the only time Beverage 2 decreased urine output compared to water was at 120 minutes(16). In the current study, cumulative urine volume was significantly decreased by Beverage 1 compared to water-alone at 120, 240, and 360 minutes (Figure 2b). Cumulative urine volume was significantly decreased with Beverage 1 compared to Beverage 2 at 60, 120, 240 and 360 minutes (Figure 2b). Bar graphs revealed that after consuming 1L of beverage urine cumulative urine volume at 360 minutes for water was 1.05L ± SD 0.32, for Beverage 1 it was 0.72L ± SD 0.28, and for Beverage 2 it was 0.97L ± SD 0.20.

**Figure 2:**
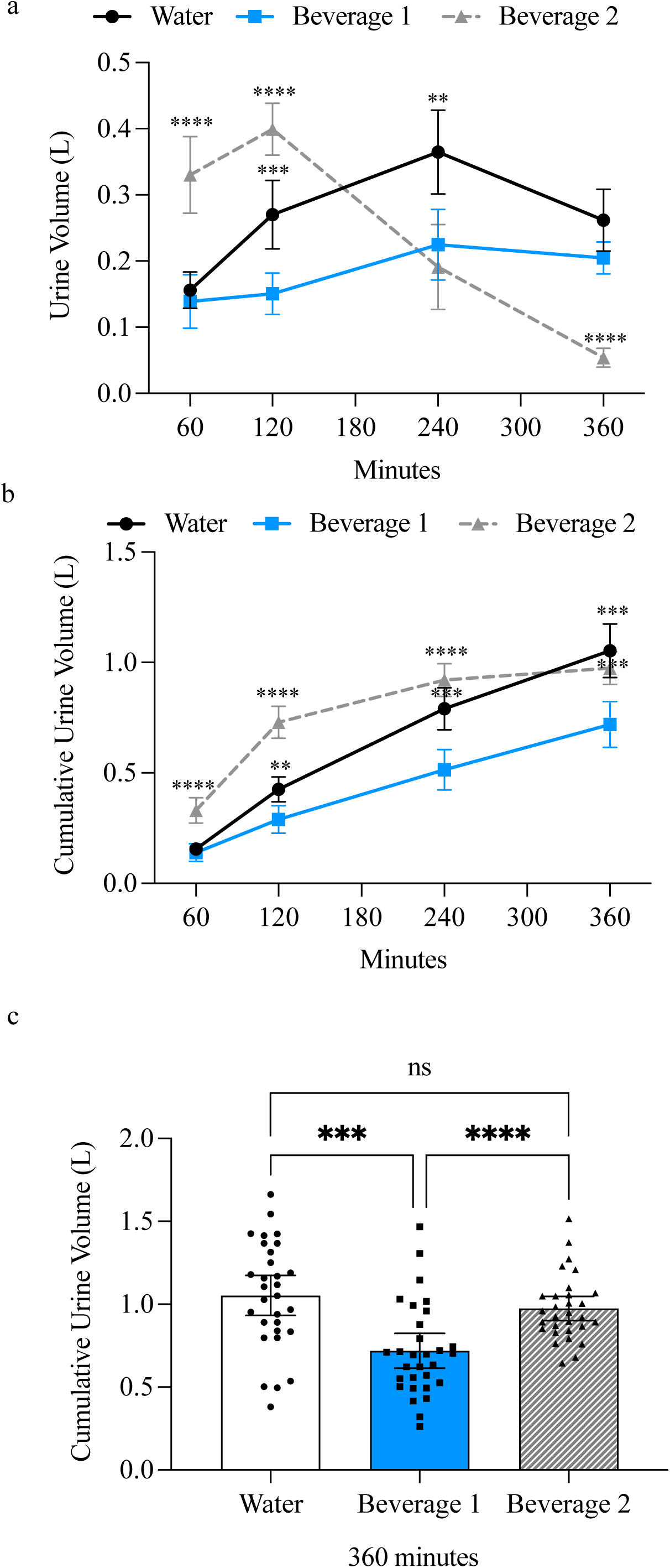
Beverage 1 significantly decreases urine output. a) Amount of urine collected at each timepoint. b) Cumulative urine output at each timepoint. For timecourse experiments error bars represent 95% confidence intervals (CI) and significance is determined by two-way Analysis of the Variance (ANOVA) at each timepoint with Tukey’s post-hoc test. **P ≤ 0.01, ***P ≤ 0.001, ****P ≤ 0.0001 comparing whether water-alone or Beverage 2 is significantly different from Beverage 1. c) Bar graph showing individual participants’ cumulative urine output after 360 minutes. Error bars represent 95% CI. Each individual value is graphed to demonstrate standard deviation between beverages. For bar graph experiments significance is determined by one-way ANOVA with Tukey’s post-hoc test. ***P ≤ 0.001, ****P ≤ 0.0001. Abbreviations: L = litre.

### Beverage 1 (Buoy) significantly increased net fluid balance

Two-way ANOVA revealed both a time-effect (P <0.0001) and beverage-effect (P <0.0001), and a time x beverage effect (P < 0.0001). The rapid ingestion (1L/30 minutes) of Beverage 2 maintained positive fluid balance until 360 minutes when it approached zero. The slow ingestion (1L/240 minutes) of water-alone maintained positive fluid balance until 360 minutes when it caused a negative fluid balance, and the slow ingestion of Beverage 1 maintained a large positive fluid balance beyond 360 minutes (Figure 3a). By 360 minutes, water-alone had a fluid balance of - 0.05L ± SD 0.32, Beverage 1 had a fluid balance of +0.28 ± SD 0.28, and Beverage 2 had a fluid balance of +0.03 ± SD 0.20. Thus, slowly-ingested Beverage 1 was at least 10-fold more effective at maintaining a positive fluid balance compared to slowly-ingested water-alone or rapidly ingested Beverage 2.

**Figure 3:**
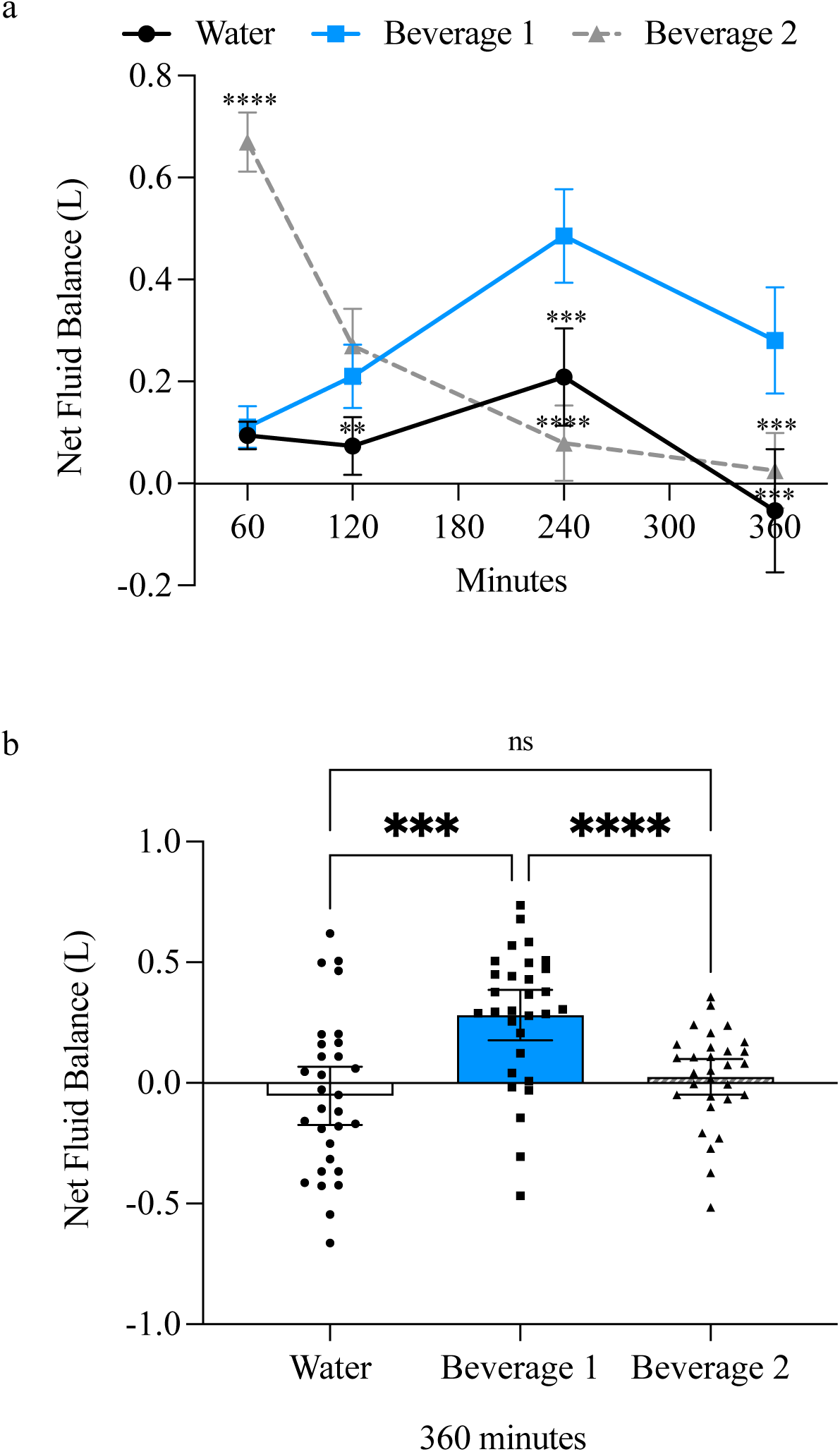
Beverage 1 significantly increases positive fluid balance. a) Net fluid balance was calculated as the amount of fluid ingested minus the cumulative urine output at each time point. For timecourse experiments error bars represent 95% confidence intervals (CI) and significance is determined by two-way Analysis of the Variance (ANOVA) at each timepoint with Tukey’s post- hoc test. **P ≤ 0.01, ***P ≤ 0.001, ****P ≤ 0.0001 comparing whether water-alone or Beverage 2 is significantly different from Beverage 1. b) Bar graph showing net fluid balance of individual participants after 360 minutes. Values > 0 are considered to represent positive fluid balance and values < 0 are considered to represent negative fluid balance. Each individual value is graphed to demonstrate standard deviation between beverages. Error bars represent 95% CI. For bar graph experiments significance is determined by one-way ANOVA with Tukey’s post-hoc test. ***P ≤ 0.001, ****P ≤ 0.0001. Abbreviations: L = litre.

### Beverage 1 (Buoy) significantly increased the beverage hydration index

Two-way ANOVA revealed both a beverage-effect (P <0.0001) and a time x beverage effect (P < 0.047), but not a time-alone effect (P = 0.077) (Figure 4a). Beverage 1 had a BHI that was significantly greater than water-alone (water = 1.0), with Beverage 1 having a value of 1.51 ± SD 0.93 at 60 minutes, 1.71 ± SD 0.81 at 120 minutes, 1.87 ± SD 1.10 at 240 minutes, and 1.64 ± SD 0.75 at 360 minutes (Figure 4a). At 360 minutes, the BHI of Beverage 1 (1.64 ± SD 0.75) was significantly higher than the BHI of bolused Beverage 2 (1.11 ± SD 0.37) despite equal amounts of sodium and volume consumed (Figure 4b). Next, BHI was analyzed based on self-reported sex. The trends were similar for both males and females and were similar to the combined results. However, only the female results reached significance. This was likely due to the small sample size, as males had an N = 14 and females had an N = 16, the power analysis estimated a sample size of 27 was required. Previous studies report no difference in BHI based on sex or body mass, thus the combined male and female data should be appropriate for statistical analysis(15, 18).

**Figure 4:**
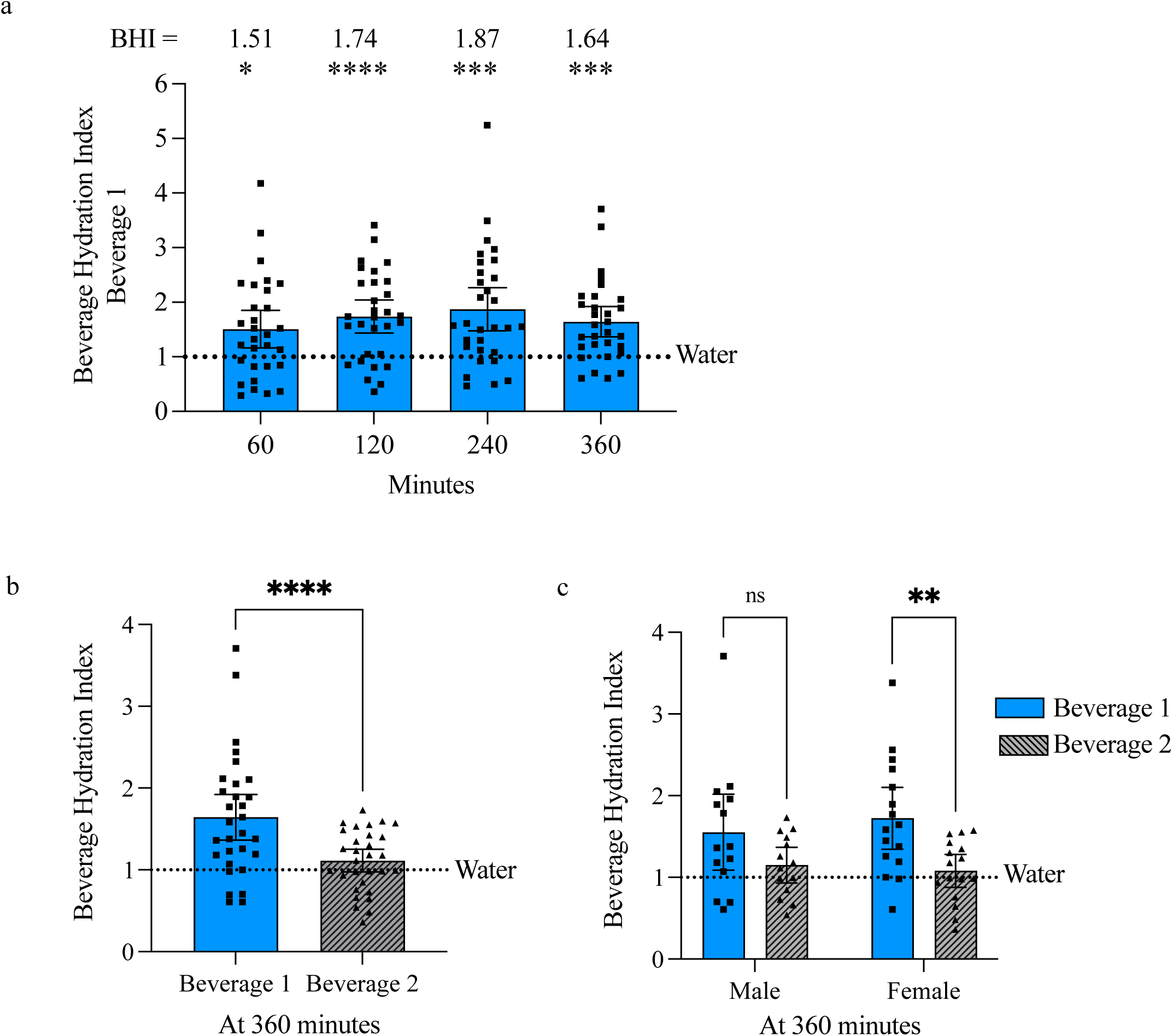
Beverage 1 significantly increases the beverage hydration index (BHI). a) BHI was calculated as the cumulative urine mass with water-alone divided by the cumulative urine mass with the intervention beverage. At each timepoint evaluated Beverage 1 significantly increases the BHI compared to water-alone. Water has a BHI set to 1.0 (shown by dotted line) and Beverage 1 has a BHI range of 1.51 to 1.87. Error bars represent 95% confidence intervals (CI) and significance is determined by two-way Analysis of the Variance (ANOVA) at each timepoint with Sidak’s multiple comparisons test. *P ≤ 0.05, **P ≤ 0.01, ***P ≤ 0.001, ****P ≤ 0.0001 comparing whether Beverage 1 is significantly different from water-alone. b) Results show calculated BHI of Beverage 1 (1.64 ± SD 0.75) compared to Beverage 2 (1.11 ± SD 0.37) at 360 mins. Error bars represent 95% CI and significance is determined by paired t test of Beverage 1 versus Beverage 2. ****P ≤ 0.0001. c) Results are grouped by subject-reported sex, the trend was the same for Beverage 1 to have a higher BHI compared to Beverage 2 when separated by sex, but only females reached a significance difference. Error bars represent 95% CI and significance is determined by two-way ANOVA at each timepoint with Uncorrected Fishers LSD post-test. **P ≤ 0.01.

### Beverage 1 (Buoy) increased cumulative urine osmolarity

Two-way ANOVA revealed both a time-effect (P <0.0001) and beverage-effect (P <0.0001), and a time x beverage effect (P < 0.0001) (Fig 5b). After ingesting 0.25 L of water-alone or Beverage 1 over 60 minutes, urine osmoles (Osms) were 658 milliOsm/kilogram (mOsm/kg) ± SD 180 for water and 764 mOsm/kg ± SD 230 for Beverage 1. Whereas initial urine Osms after ingesting 1L Beverage 2 over 30 minutes were 343 mOsm/kg ± SD 251. Over the next 360 minutes the water-alone and Beverage 1 similarly decreased urine Osms to a nadir of ∼330 mOsms/kg, whereas Beverage 2 decreased to a nadir of ∼140 mOsm/kg at 120 minutes and then increased to 789 mOsm/kg by 360 minutes (Figure 5a). The change in urine Osms at each timepoint likely reflected the rate at which the beverage was consumed. When cumulative urine Osms were evaluated at 360 minutes there was no significant difference between Beverage 1 (1928 mOsm/kg ± SD 544) and Beverage 2 (1866 mOsm/kg ± SD 318), however the urine Osms of water (1666 mOsm/kg ± SD 454) were significantly lower compared to Beverage 1 (Figure 5b and c).

**Figure 5:**
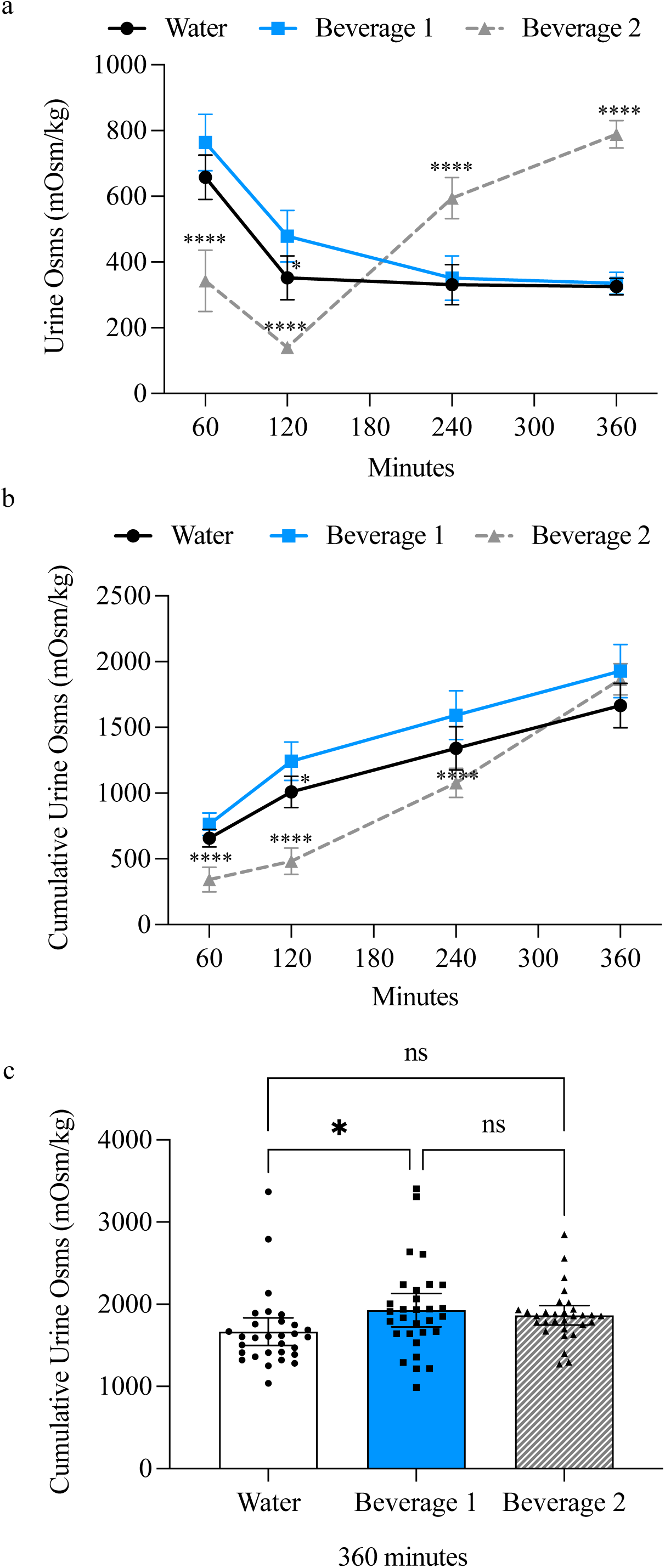
Beverage 1 significantly increases cumulative urine osmolarity (osms) compared to water-alone. a) Timecourse of urine osms and b) Cumulative urine osms. Error bars represent 95% confidence intervals (CI) and significance is determined by two-way Analysis of the Variance (ANOVA) at each timepoint with Tukey’s multiple comparisons test. *P ≤ 0.05, ****P ≤ 0.0001 comparing whether Beverage 2 or water-alone is significantly different from Beverage 1. c) Bar graphs showing individual participants’ cumulative urine osms at 360 minutes after consuming all three beverages. There is a significant increase in urine osms in the Beverage 1 group [1928 mOsm/kilogram (kg) ± SD 544] compared to water-alone (1666 mOsm/kg ± SD 454). Beverage 2 is not significantly different from either beverage (1866 mOsm/kg ± SD 318). Error bars represent 95% CI. Each individual value is graphed to demonstrate standard deviation between beverages. For bar graphs significance is determined by one-way ANOVA with Tukey’s post-hoc test. *P ≤ 0.05. Abbreviations: Osms = osmolarity. Kg = kilogram. Ns = not significant

#### Beverage 1 (Buoy) decreased cumulative urine sodium and chloride compared to water-alone

The concentration milliequivalent/L (meq/L) of urine Na+, K+, and Cl- followed a similar trend to urine Osms, with both water-alone or Beverage 1 initially decreasing the electrolyte concentration in the urine and then plateauing after 120 minutes, whereas Beverage 2 caused an initial decrease that began to rise after 120 minutes (Figure 6a-c). The urine electrolyte concentration was dependent on the rate of beverage ingestion and the volume of urine output. Therefore, to normalize for urine dilution and/or concentration, the urine electrolyte concentrations were normalized to either urine creatinine (Figure 6d-f) or urine volume (Figure 6g-i). Both normalization methods showed a similar trend, that the excretion of urine electrolytes (meq) tended to be higher with water-alone compared to Beverage 1, most notably at 240 minutes. Cumulative urine Na+ concentration (meq) was consistently higher with water-alone compared to Beverage 1 at 120, 240, and 360 minutes (Figure 6j & m). Cumulative urine K+ (meq) was only different between water-alone and Beverage 2 at 360 minutes (Figure 6k & n).

**Figure 6:**
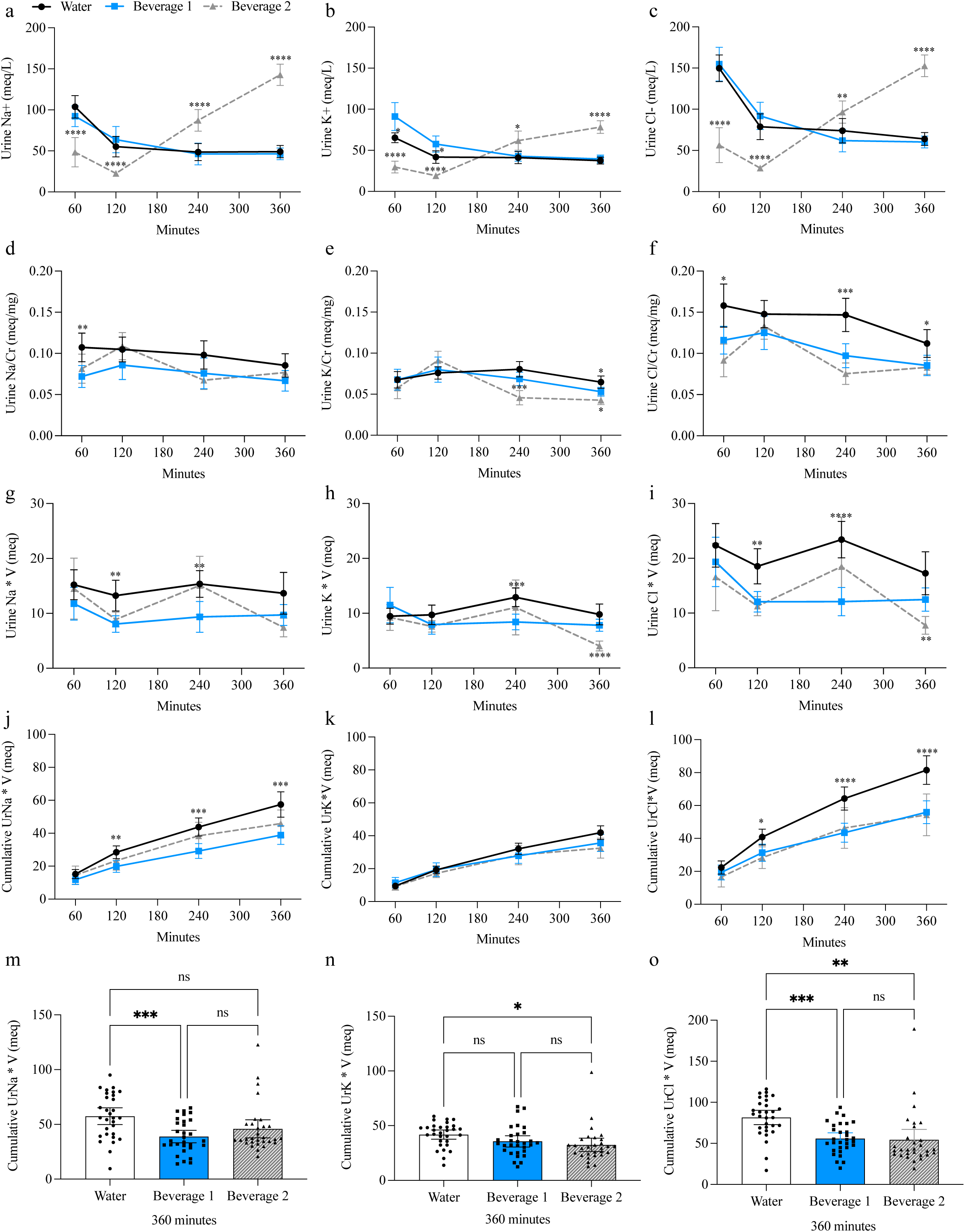
**Beverage 1 significantly decreases cumulative urine electrolytes compared to water-alone.** a-c) Timecourse of urine electrolyte concentration. d-f) Timecourse of urine electrolytes normalized to urine creatinine. g-i) Timecourse of urine electrolytes normalized to urine volume. j-l) Cumulative timecourse of urine electrolytes normalized to urine volume for total electrolytes excreted in the urine over 360 minutes For timecourse experiments error bars represent 95% confidence intervals (CI) and significance is determined by two-way Analysis of the Variance (ANOVA) at each timepoint with Tukey’s multiple comparisons test. *P ≤ 0.05, **P ≤ 0.01, ***P ≤ 0.001, ****P ≤ 0.0001 comparing whether Beverage 2 or water-alone is significantly different from Beverage 1. m-o) Bar graphs showing individual participants’ cumulative urine electrolytes normalized for volume at 360 minutes after consuming all three beverages. Beverage 1 had significantly less sodium (Na+) (39 meq ± SD 15) and chloride (Cl-) (56 meq ± SD 19) compared to water- alone Na+ (58 meq ± SD 21) and Cl- (82 meq ± SD 23). Error bars represent 95% CI. Each individual value is shown to demonstrate standard deviation between beverages. For bar graphs significance is determined by one-way ANOVA with Tukey’s post-hoc test. *P ≤ 0.05, ***P ≤ 0.001. One value for urine potassium (K+) and urine Cl- were below the limit of detection and excluded from results. Abbreviations: Na+ = sodium. K+ = potassium. Cl- = chloride. UrNa = urine sodium. UrK = urine potassium. UrCl = urine chloride. V = volume. Meq = milliequivalents. Mg = milligrams. L = litres.

Whereas cumulative urine Cl- (meq) was significantly less for Beverage 1 and Beverage 2 compared to water-alone (Figure 6l & o). Beverage 1 had significantly less sodium (Na+) (39 meq ± SD 15) and chloride Cl- (56 meq ± SD 19) compared to water-alone Na+ (58 meq ± SD 21) and Cl- (82 meq ± SD 23).

## DISCUSSION

The importance of hydration is often underappreciated and under-researched, despite water being the largest single constituent of the human body and the most essential nutrient for survival(4, 19, 20). In this current study we have shown that the ingestion of Beverage 1 (Buoy) improved hydration status in participants at rest based on several markers of hydration including decreased urine volume, positive net fluid balance, and increased beverage hydration index (BHI)(8). After six hours Beverage 1 had a 64% increase in the BHI compared to water-alone and a 48% increase in BHI compared to Beverage 2. The BHI is a relatively new tool to measure the hydration index(8). Since its inception in 2016, it has been adapted for several studies to compare the hydrating capabilities beverages ranging from water, milk, coffee, sports beverages, and oral rehydration solutions(8, 15–18, 21). BHI does not appear to be affected by body mass or sex(18), but is affected by advanced age(17). For that reason, we limited the age of the study participates to 18-45 years, and collectively analyzed the results regardless or sex or body mass.

It has long been known that ingestion of plain water is ineffective at maintaining euhydration due to reductions in plasma osmolality that induce prompt diuresis(22). In our own study we found that water-alone increased urinary electrolyte excretion (meq). The increase in cumulative urine electrolyte excretion with water-alone may seem counterintuitive since the participants consumed less Na+, K+, and Cl- with water-alone, but this study highlights how water-induced diuresis can be disadvantageous to hydration and increase the risk of electrolyte derangements. The addition of electrolytes, protein and carbohydrates to water can augment fluid retention and improve hydration. Prior studies have shown that drinks containing the highest macronutrient and electrolyte content are the most effective at increasing BHI—with the BHI 2 hours after bolused skim milk, oral rehydration solution or orange juice being 1.58, 1.54, 1.39, respectively compared to water-alone(8). Prior studies have also shown a trend for rapidly ingested Beverage 2 (1L/30minutes) to have an increased BHI of 1.2 at 2 hours, although it did not reach statistical significance when analyzed via ANOVA(16).

Sodium is the most common electrolyte studied in hydration beverages. Research has shown that to maximize hydration the sodium concentration should typically be between 40-100 millimole (mmol)/L (920-2300 mg/L), whereas drinks with lower sodium concentrations of 20-30 mmol/L (460-690 mg/L) have inconsistent results and do not always improve markers of hydration(8, 17, 18, 21). This is important, because most commercially available sports drinks have a sodium concentration ranging from 20-30 mmol/L. Compare this to Pedialyte which is closer to 45 mmol/L, or pharmaceutical oral rehydration solutions (ORS) that have a sodium concentration > 75 mmol/L(15, 21). A key issue with oral rehydration solutions containing higher sodium contents is that they are unpalatable due to the salty flavor. For this current study we chose a sodium concentration of 26 mmol/L (600 mg/L). Although this is within the lower sodium range, this dose was chosen because it is similar to prior studies looking at the hydration efficacy of Beverage 2 (16), it is closer to the actual daily amount recommended, and avoids the unpalatable flavor of higher sodium doses.

Despite prior studies showing inconsistent results with sodium concentrations between 20-30 mmol/L(8, 17, 18, 21), this current study found that Beverage 1 containing 26 mmo/L (600mg/L) of sodium significantly decreased urine volume, increased net fluid balance, improved the BHI, and increased urine Osms compared to water-alone. Moreover, Beverage 1 was more effective at improving markers of hydration than Beverage 2, despite ingestion of identical amounts of fluid (1L) and sodium (600 mg). There were several factors that may explain these differences including rate of ingestion, carbohydrate content, and chloride concentration.

Prior studies with rapid ingestion rates of 1L/30 minutes demonstrated that negative fluid balance with water typically occurs within 120 minutes whereas electrolyte-containing beverages extend the time in positive fluid balance(8, 15, 16). Only a few studies have evaluated the impact of ingestion rate on hydration status, among these studies there is a consensus that slower “metered” rates of ingestion are likely to result in more efficient hydration compared to “bolus” ingestion(6, 23–25). This is due to the protective *bolus response* that occurs with the rapid ingestion of hypotonic fluid(26). The hypotonic fluid reduces plasma osmolarity, and to protect against hyponatremia the body inhibits arginine vasopressin (AVP) release resulting in a prompt diuresis. Despite the paucity of data, it seems clear that large boluses of fluid should be avoided—rather beverages should be consumed over several hours for effective hydration(6).

Traditionally, BHI is measured after the rapid ingestion of 1L bolus during the first 30 minutes of the study (7, 15, 17, 22, 23). To our knowledge only one other study measured BHI with slower fluid ingestion over hours(25). Thus, as a positive control we included Beverage 2 with rapid ingestion (1L/ 30 minutes) as it has previously been shown to improve markers of hydration(16). For this study, Beverage 1 was administered as a slower ingestion over four hours and then compared to the hydration efficacy of water-alone also given over four hours or Beverage 2 given as a bolus. We chose to study Beverage 1 over a slower time course because the manufacturer’s label recommends consuming the beverage in multiple servings throughout the day. Based on the BHI after six hours, our results suggest that Beverage 1 with “metered” ingestion is superior to “metered” water ingestion or to Beverage 2 with “bolus” ingestion, however a key limitation in evaluating rate of ingestion is that we did not directly compared Beverage 1 as a bolus ingestion. Evaluating rate of ingestion remains an important area of hydration research. Beyond ingestion rate, there are other variables within the beverages that could affect hydration including macronutrients and electrolytes.

It is common for sports drinks and oral rehydration beverages to have differing compositions of carbohydrates, amino acids and electrolyte content. For hydration efficacy, the electrolyte content appears to be the most important factor, but carbohydrates and amino acids also play a role(6, 15, 17, 21). Prior studies have shown that the addition of carbohydrates to beverages can enhance hydration by prolonging gastric emptying and by stimulating glucose-sodium co-transporters that promote an osmotic gradient to facilitate water absorption via the intestinal tract(21, 27–29). However, the carbohydrate concentration must be greater than what is typically found in sports drinks to consistently improve hydration(21). Moreover, the addition of calories, carbohydrates, and artificial flavoring can be a barrier for people who need to avoid excess calories or carbohydrates. Furthermore, artificial sweeteners such as sorbitol have actually been shown to cause diuresis(30). For this current study, Beverage 1 was carbohydrate-free and artificial sweetener free, whereas Beverage 2 contained 2 grams of sugar and 8 grams of carbohydrates as well as sorbitol.

Potassium levels were also different among the beverages in this study, with it being more than two-fold higher in Beverage 2 compared to Beverage 1. This is unlikely to have influenced hydration as potassium has not been shown to enhance fluid retention during post-exercise rehydration when sodium is adequate(6). A striking difference in this study is that Beverage 1 contained over ten-fold more chloride compared to Beverage 2 (Beverage 1 had 960 mg or 27mmol/L compared Beverage 2 with 80mg or 2.3 mmol/L). Chloride is the most abundant anion in the body and is critical for maintaining osmotic pressure, acid-base balance, the movement of water, and chloride is responsible for about a 1/3 of extracellular fluid tonicity(31). Others have postulated that chloride might be important for hydration given that it is often the anion associated with sodium, however the role of chloride remains obscure compared to its sodium counterpart (6, 31). To our knowledge there are no studies addressing the efficacy of chloride in improving hydration compared to alternative anions, such as bicarbonate or citrate (6, 29, 32). In commercial sports drinks, bicarbonate and citrate are sometimes substituted for chloride to reduce carbohydrate induced GI symptoms, to buffer changes to blood pH during exercise, and to improve palatability(29). Future studies should focus on isolating the effects of chloride independent from sodium and compare the hydration efficacy to other anions.

A limitation to this study is that we did not power the study to differentiate between males and females even though there are sex-based differences in water handling. Total body water and water intake varies between males and females, with females having lower total body water and total water intake(4). This is attributed to females having lower metabolic rates, smaller size, and lower fat-free mass(4). Additionally, males tend to sweat more and have higher electrolyte losses(3). Males have been found to be more sensitive to AVP which inhibits diuresis, whereas females tend to have more estrogen and progesterone that enhance water and electrolyte retention(3). Despite these differences the American College of Sports states that “sex differences in renal water and electrolyte retention are subtle and probably not of consequence”(3). Reassuringly, previous studies support that BHI measurements are not affected by sex(18). When we analyzed our BHI results by sex, the trends between males and females remained the same. However, only the female BHI results reached significance, likely due to the results being underpowered for individual sexes.

While the field of hydration research is growing, in many ways it remains in its infancy and many questions remain unanswered. Much of the research has focused on the acute effects of hypohydration on athletes and physical performance, yet the implications for hydration extend into overall health outcomes(7, 19, 33). The United States Health Care industry spends billions of dollars a year treating the preventable condition of dehydration(1, 34, 35). Chronic hypohydration has been associated with increased morbidity and mortality including headaches, kidney stones, exercise-related asthma, heart disease, kidney disease, blood clots, stroke, dental disease, urinary tract infections, bladder and colon cancer, gallstones, mitral valve prolapse, and glaucoma(4, 7, 33).

Future studies will be aimed at understanding how Beverage 1 (Buoy) may benefit populations extending beyond healthy, fit participants, and will include studying vulnerable populations including the elderly(7, 17), those with GI disease(36), and cancer patients(37, 38). More studies are needed to understand how ingestion rate and beverage composition affect beverage hydration efficacy. Hydration is essential to our survival, thus, there is a need for more research focused on optimizing hydration to improve performance, health promotion and disease prevention.

## Data Availability

All data produced in the present study are available upon reasonable request to the authors.

## CONTRIBUTORS

### Contributors

All authors read and approved the final manuscript. CBS, ER, HS, AMonroe designed and implemented the program. CBS, ER, NZ collected, analyzed and verified data. CBS performed the statistical analysis and interpreted results. AMahajan designed the program. All authors read and critically reviewed and revised the manuscript.

### Data Sharing Statement

The datasets generated and/or analysed during the current study are not publicly available but are available from the corresponding author on reasonable request via email, and will be shared via secure platform, after approval by corresponding author, with a signed data access agreement.

### Ethics approval and consent to participate

Institutional Review Board (IRB) at the University of Pittsburgh approved this study with a waiver of consent (STUDY22090018).

### Funding

UPMC Department of Anesthesiology and Perioperative Medicine

